# INTERNATIONAL PREVALENCE OF CONSULTATION WITH A NATUROPATHIC PRACTITIONER: A SYSTEMATIC REVIEW AND META-ANALYSIS

**DOI:** 10.1101/2021.08.08.21261774

**Authors:** Amie Steel, Rebecca Redmond, Janet Schloss, Holger Cramer, Joshua Goldenberg, Matthew Leach, Joanna Harnett, Claudine Van de Venter, Andy McLintock, Ryan Bradley, Jason Hawrelak, Kieran Cooley, Brenda Leung, Jon Adams, Jon Wardle

## Abstract

**Objectives:** Naturopathy is a traditional medicine system informed by codified philosophies and principles, and an emphasis on non-pharmacologic therapeutic interventions. While naturopathy is practiced by approximately 75 000 to 100 000 naturopathic practitioners in at least 98 countries, little is known about the international prevalence of history of consultation with a naturopathic practitioner. This study reports a systematic review and meta-analysis of studies describing the global prevalence of history of consultation with a naturopathic practitioner by the general population.

**Setting:** The included literature was identified through a systematic search of eight databases between September and October 2019, as well as the grey literature.

**Participants:** Studies were included if they reported the prevalence rate of consultations with a naturopathic practitioner by the general population

**Interventions:** Survey items needed to report consultations with a naturopathic practitioner as defined in the country where data was collected, and not combine naturopathic consultations with other health services or only report consulations for illness populations.

**Primary and secondary outcome measures:** Primary measures used for the analysis was consultations in the previous 12-months. Other prevalence timeframes were reported as secondary measures.

**Methods:** Meta-analysis of prevalence data was conducted using random effects models based on individual countries and World Health Organisation (WHO) world regions.

**Results:** The literature search identified eight manuscripts summarizing 13 studies reporting prevalence for inclusion in the review. All included studies had a low risk of bias. Meta-analysis of the included studies by world region found the 12-month prevalence of history of naturopathy consultations ranged from 1% in the Region of the Americas to 6% in the European and Western Pacific Regions.

**Conclusions:** There are up to 6-fold differences in the prevalence of naturopathy consults over 12-months between and within world regions, which may be driven by a range of policy, legislative and social factors.

**Strengths and Limitations of this study:**

- Naturopathy is one of the most commonly used traditional and complementary medicines in the Western world and this is the first systematic review and meta-analysis reporting the prevalence of consutations with a naturopathic practitioner.
- This study includes only includes data published after 2010 to ensure the results are contemporary, however this may have excluded some studies in countries with older data.
- The results are limited by the poor availability of data reporting consultations with a naturopathic practitioner, including in countries where a large number of naturopathic practitioners are known to provide care.

## Introduction

Naturopathy is a traditional medicine system underpinned by six philosophical principles (see Table 1), which were codified by the profession in the 20^th^ century [1]. These philosophical principles characterize naturopathic practice and are globally accepted by the profession [2]. Other defining tenets of naturopathic practice are patient-centeredness and individualization, with naturopaths typically drawing upon a range of therapeutic interventions (e.g., diet and lifestyle counselling, herbal medicine, nutritional supplementation, manual therapies, and mind-body practices) to best meet the health care needs and preferences of the patient [3]. Globally, naturopathy is practiced in at least 98 countries with representation in every world region [4]. Naturopathy is practiced widely in Europe (n=54 practicing countries), followed by Latin America (n=51), Africa (n=47), and the Western Pacific (n=37) [5]. Estimates from the World Naturopathic Federation suggest there are between 75,000 and 100,000 naturopaths currently in clinical practice across the world [5].

**Table 1.**
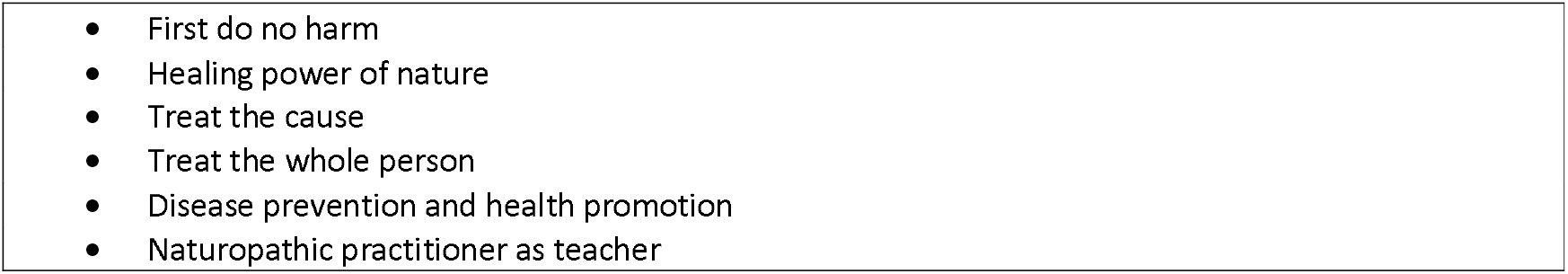
Philosophical principles of naturopathy [3]

Training of the naturopathic workforce is currently provided by an estimated 90 education institutions globally, with entry-level qualifications ranging from technical diploma to clinical doctorate [3]. The curriculum of these naturopathic programs typically includes content in health sciences (e.g., anatomy, physiology, chemistry, and biochemistry), clinical sciences (e.g. clinical examination, differential diagnosis), social sciences (e.g. psychology, counselling), and naturopathic sciences (e.g. nutritional medicine, herbal medicine, lifestyle medicine, dietary modification, homeopathy, and manual therapies) [4]. Despite similarities in the content of these training programs, naturopathic scope of practice varies considerably across jurisdictions due to differences in regulation and legislative requirements [6].

In response to an increase in the use of traditional and complementary medicine (including the utilization of naturopathic health services), the World Health Organisation has developed global strategies to ensure access to safe and effective healthcare, which include promoting the integration of traditional and complementary therapies (including naturopathy) into healthcare systems [7]. Several international research studies suggest the demand for naturopathic services may be attributed to personal healthcare beliefs, dissatisfaction with biomedical care, increased disease severity, and unmet healthcare needs [8-15]. Nevertheless, the global use of naturopathic services is not well understood. Therefore the aim of this study was to describe the prevalance of a history of consultations with naturopathic practitioners globally, including potential differences across world regions.

## Methods

### Aim

This study aims to describe the global prevalence of a history of consultation with a naturopathic practitioner by the general population.

### Study Design

A systematic review and meta-analysis of prevalence studies were undertaken in accordance with the AMSTAR 2 guidelines [16]. The protocol for this review was submitted to PROSPERO on the 2nd September, 2019 and was registered on the 28th April, 2020 [CRD42020145529].

### Inclusion and exclusion criteria

Articles were included that reported original data from cohort studies, cross-sectional studies, survey research, case-control studies, prevalence studies, or epidemiologic studies. Studies reporting on the general population prevalence of consultations with a naturopathic practitioner either in the previous 12 months or over the user’s lifetime were considered for inclusion. All relevant papers were included irrespective of language of publication or risk of bias score. Articles were excluded that presented results from specific sub-patient populations (e.g. children, female or male specific, age limitations, illness populations). Studies were also excluded if they only presented the prevalence of consultations with other health professionals that may use treatments commonly associated with naturopathy (e.g. herbal medicine, hydrotherapy, yoga, etc) but were not explicitly named as naturopathic practitioners, or where naturopathic consultation rates were conflated with a cumulative group of health services (such as complementary and alternative medicine [CAM]). To ensure the analysis reflected contemporary patterns of use, studies were excluded if they were published before 2010.

### Search Strategy

A systematic electronic search of the following databases was conducted between 6th September and 2^nd^ October 2019: MEDLINE, AMED, EMBASE, CINAHL, Global Health, WHO Iris, PROQUEST dissertations database, and Lilac. The complete search strategy for MEDLINE is presented in Table 2. A search for grey literature was also performed. The search targeted countries where, according to the WHO Global Report on Traditional and Complementary Medicine (2019) [20], naturopathic practitioners provide care to the community. The search was performed using the Google search engine and the terms *prevalence, use, naturopathy, report*, and the country name.

**Table 2:**
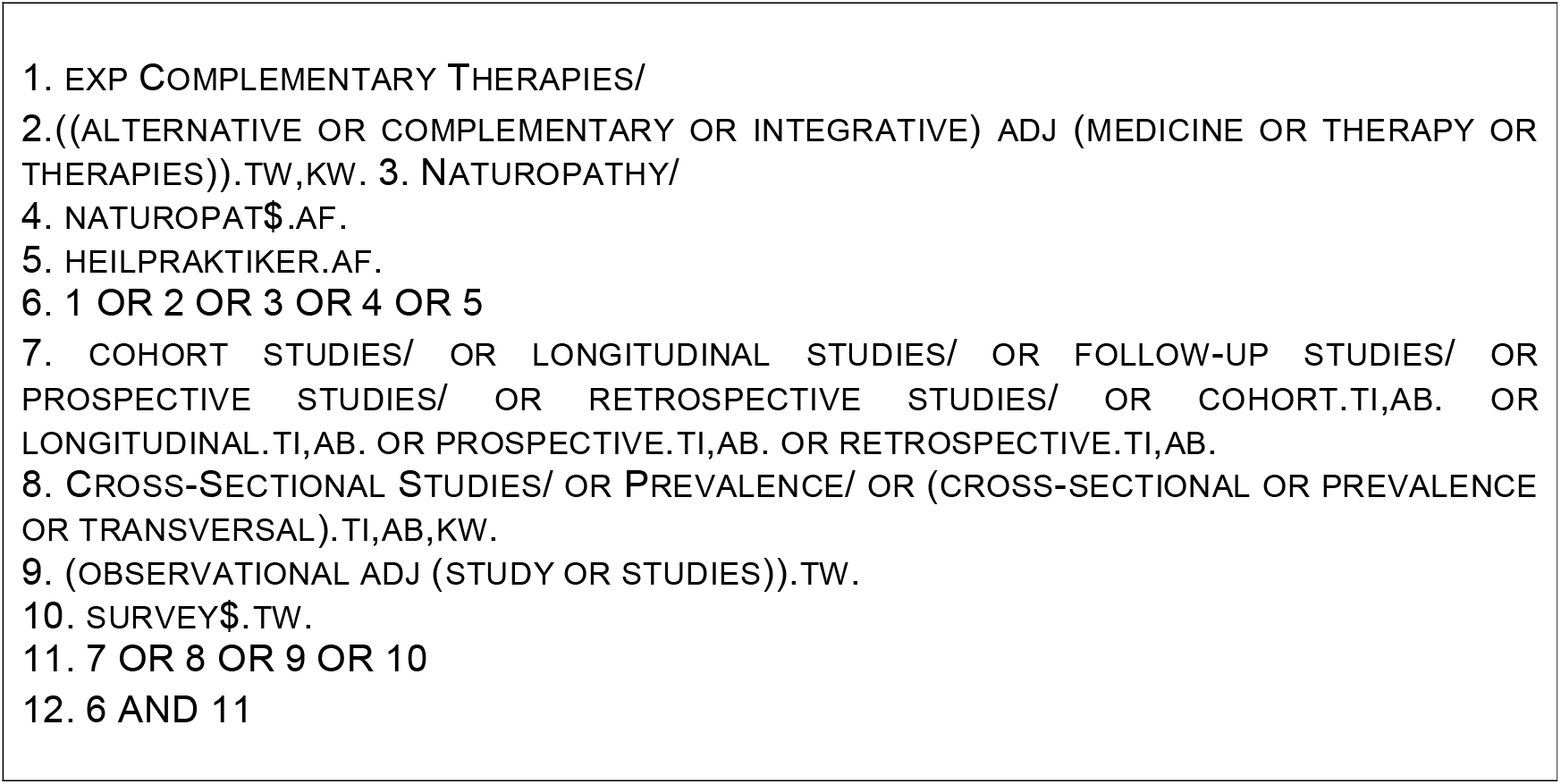
Example search terms applied to database searches

### ARTICLE IDENTIFICATION AND SELECTION

A list of all citations identified through the search were exported from each database by AM and uploaded to Covidence [17] for filtering and selection. Initial screening of title and abstracts against the inclusion/exclusion criteria was conducted by AM. Two members of the authorship team (AM and AS) then independently reviewed the full text of the remaining citations to determine their suitability against the same criteria. Any differences were resolved through discussion between both reviewing authors. The list of bibliographic references and subsequent citations (identified through Google Scholar) of included papers were also checked by AS to identify additional articles otherwise missed through the database search. JHar and JS extracted data from the included papers. AS and JS assessed the papers for quality of reporting against the STROBE checklist [18]; risk of bias was assessed using the tool developed by Hoy et al [19] by JG and JAH. Differences in scoring for both tools were resolved through discussion until consensus was achieved.

### Analysis

The results were grouped for narrative presentation of results in accordance with the World Health Organisation (WHO) world regions [21]. Where studies reported the results of more than one year, these were treated as different studies in the analysis. Articles with unclear numerators or denominators were calculated by the research team where the necessary information was provided or checked against source documents for the same study. Authors were contacted to verify information not able to be determined through these other methods.

Prevalence rates and standard errors were calculated using a standardized Microsoft Excel (version 12.3.5, Microsoft, Redmond, USA) spreadsheet [22]. Review Manager software (version 5.3, Nordic Cochrane Centre, Copenhagen, Denmark) was used to conduct the meta-analysis, using random effects models by the Generic Inverse Variance method. Weighted prevalence rates with 95% confidence intervals (95% CI) were calculated for 12-month prevalence and lifetime prevalence separately. Separate analyses were conducted for a) country of origin and b) WHO world regions.

Heterogeneity between studies was estimated on the basis of the raw proportions, by using the I^2^ statistic. Intervals were defined as per published guidance [23, 24]: low heterogeneity (I^2^ of 0–24%); moderate heterogeneity (I^2^ of 25–49%); substantial heterogeneity (I^2^ of 50–74%); relevant heterogeneity (I^2^ of 75–100%). In order to assess heterogeneity, χ^2^ tests were conducted with p ≤ 0.10 [24]. We intended to perform sensitivity analyses to compare differences between outcomes on all studies to studies with low risk of bias only (defined as <4 items recorded as ‘no’ on the Hoy et al tool). However, as all studies were classified as low risk of bias, this was not possible.

### Ethics approval

As this study presents a review and synthesis of published research and does not engage with data collection of human or animal subjects, it is deemed negligible risk and no ethics approval was required.

## Results

### Search characteristics

The article selection process is presented in Figure 1. The database search identified 13,968 citations including 2,509 duplicates. Of these, 11,374 were excluded through title and abstract screening. The full text of the remaining 85 articles were assessed for eligibility, of which 78 were excluded for the following reasons: not reporting naturopathic consultations (n=54), conference abstract only (n=9), not original research (n=7), wrong outcomes reported (n=5), overlooked duplicate (n=2), and wrong study design (n=1) (full list of excluded studies available in Supplementary File 1). This resulted in seven articles being retained. A search for grey literature using the Google search engine was also performed, and targeted countries where, according to the WHO Global Report on Traditional and Complementary Medicine (2019) [6], naturopaths/naturopathic doctors are providing care to the community. The reference lists and subsequent citations of the remaining articles were checked and when combined with the results of the Google Search, resulted in identification of an additional 19 articles (3 references and 16 citations), of which one report was found to meet the inclusion criteria for this review. This yielded a total of eight included studies, one of which was published in a report.

**Figure 1:**
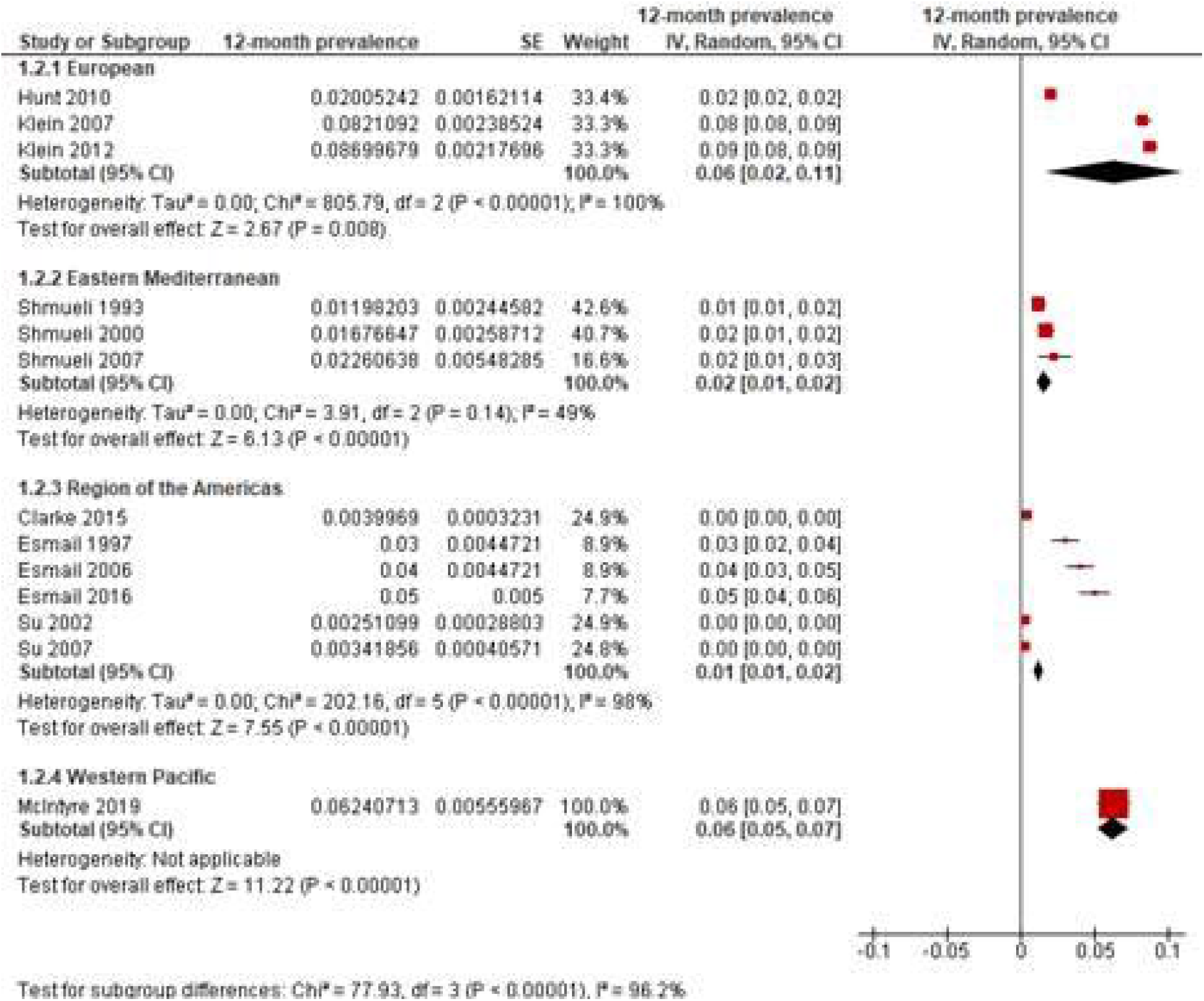
Flow chart representing article selection method in line with PRISMA protocol

### Study characteristics

The included studies reporting 12-month prevalence of naturopathy use in a national population were represented across four of the six WHO world regions: European (n=2) [25, 26], Eastern Mediterranean (n=1) [27], Region of the Americas (n=3) [28-30], and the Western Pacific (n=1) [31] (see Table 3). One of the studies from Canada presented the lifetime prevalence of naturopathy use [30], and an additional study from India (South East Asian World region) did not specify the time period during which naturopathy was used [21] (see Table 4).

**TABLE 3:**
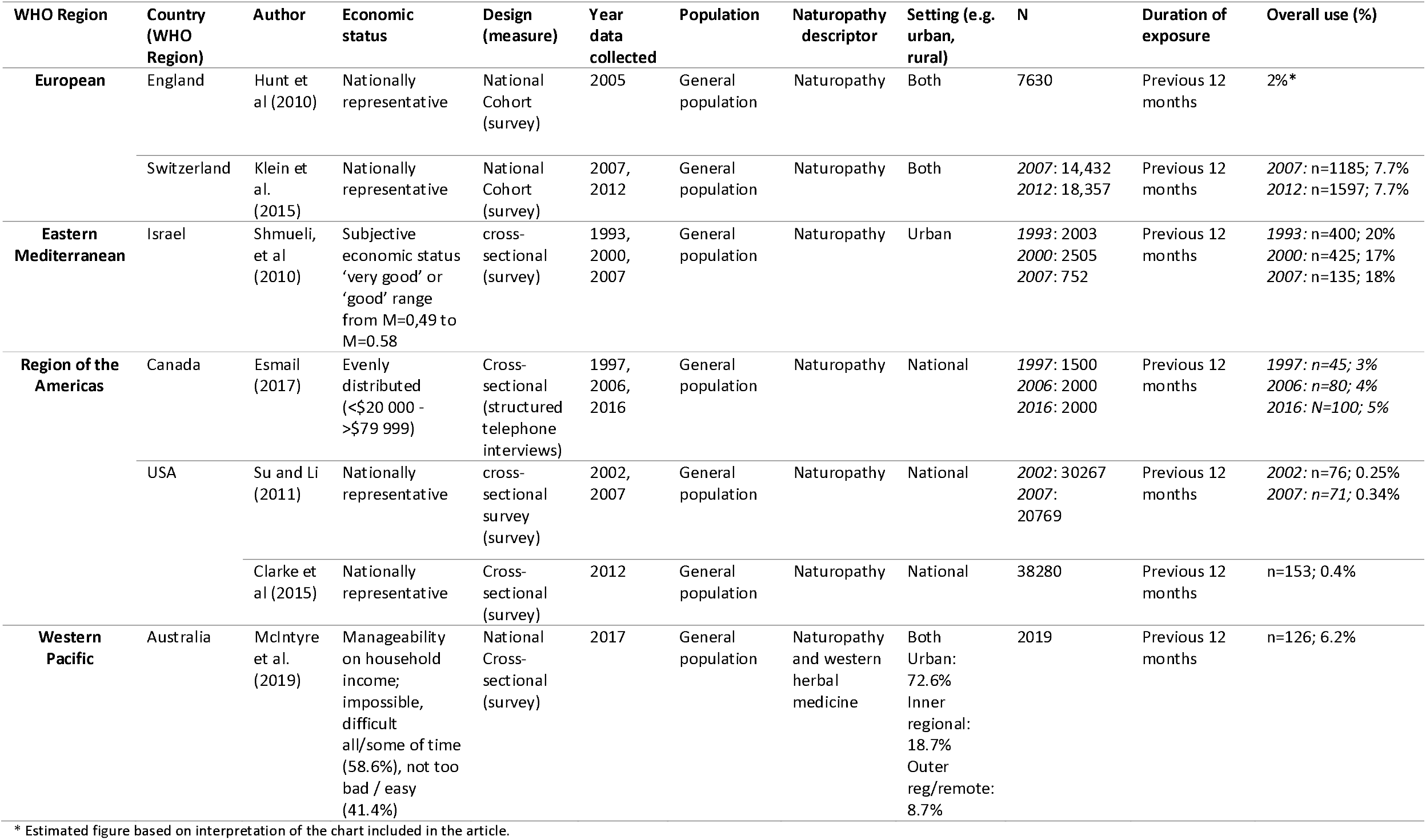
SUMMARY INFORMATION OF INCLUDED STUDIES REPORTING PREVALENCE OF USE OF NATUROPATHY IN THE PREVIOUS 12 MONTHS.

| WHO Region | Country (WHO Region) | Author | Economic status | Design (measure) | Year data collected | Population | Naturopathy descriptor | Setting (e.g. urban, rural) | N | Duration of exposure | Overall use (%) |
| --- | --- | --- | --- | --- | --- | --- | --- | --- | --- | --- | --- |
| European | England | Hunt et al (2010) | Nationally representative | National Cohort (survey) | 2005 | General population | Naturopathy | Both | 7630 | Previous 12 months | 2%* |
|  | Switzerland | Klein et al. (2015) | Nationally representative | National Cohort (survey) | 2007, 2012 | General population | Naturopathy | Both | 2007: 14,432<br>2012: 18,357 | Previous 12 months | 2007: n=1185; 7.7%<br>2012: n=1597; 7.7% |
| Eastern Mediterranean | Israel | Shmueli, et al (2010) | Subjective economic status 'very good' or 'good' range from M=0.49 to M=0.58 | cross-sectional (survey) | 1993, 2000, 2007 | General population | Naturopathy | Urban | 1993: 2003<br>2000: 2505<br>2007: 752 | Previous 12 months | 1993: n=400; 20%<br>2000: n=425; 17%<br>2007: n=135; 18% |
| Region of the Americas | Canada | Esmail (2017) | Evenly distributed (<\$20 000 - >\$79 999) | Cross-sectional (structured telephone interviews) | 1997, 2006, 2016 | General population | Naturopathy | National | 1997: 1500<br>2006: 2000<br>2016: 2000 | Previous 12 months | 1997: n=45; 3%<br>2006: n=80; 4%<br>2016: N=100; 5% |
|  | USA | Su and Li (2011) | Nationally representative | cross-sectional survey (survey) | 2002, 2007 | General population | Naturopathy | National | 2002: 30267<br>2007: 20769 | Previous 12 months | 2002: n=76; 0.25%<br>2007: n=71; 0.34% |
|  |  | Clarke et al (2015) | Nationally representative | Cross-sectional (survey) | 2012 | General population | Naturopathy | National | 38280 | Previous 12 months | n=153; 0.4% |
| Western Pacific | Australia | McIntyre et al. (2019) | Manageability on household income; impossible, difficult all/some of time (58.6%), not too bad / easy (41.4%) | National Cross-sectional (survey) | 2017 | General population | Naturopathy and western herbal medicine | Both Urban: 72.6%<br>Inner regional: 18.7%<br>Outer reg/remote: 8.7% | 2019 | Previous 12 months | n=126; 6.2% |

\* Estimated figure based on interpretation of the chart included in the article.

**TABLE 4:** SUMMARY INFORMATION OF INCLUDED STUDIES REPORTING PREVALENCE OF USE OF NATUROPATHY OVER OTHER TIME PERIODS.

| WHO Region | Country (WHO Region) | Author | Economic status | Design (measure) | Year data collected | Population | Naturopathy descriptor | Setting (e.g. urban, rural) | N | Duration of exposure | Overall use (%) |
| --- | --- | --- | --- | --- | --- | --- | --- | --- | --- | --- | --- |
| Region of the Americas | Canada | Esmail (2017) | Evenly distributed (<\$20 000 - >\$79 999) | Cross-sectional survey | 1997, 2006, 2016 | General population | Naturopathy | Both | 1500 (1997); 2000 (2006); 2000 (2016) | Ever used | 1997: 6%<br>2006: 9%<br>2016: 11% |
| South-East Asian | India | Srinivasan and Raji Sugumar (2017) | Diversity of occupation, social group, education, and religion | Cross-sectional (survey) | 2011-2012 | Households in the general population | Naturopathy and yoga | Both | Total: 65507<br>Urban: 26996<br>Rural: 38511 | Not specified | Total: n=6616 (10%)<br>Urban: n=3227 (12%)<br>Rural: n=2607 (7%) |

All included studies sampled the general adult population and reported data from a nationally representative sample or demonstrated a distribution of economic categories, except for one study from Israel whereby the majority of participants’ subjective economic status was rated as ‘very good’ or ‘good’ [27]. Four studies included prevalence data from more than one time point [26-28, 30], with the earliest data collected in 1993 [27]. Two papers reported data from the same national cohort study, but from different time points [28, 29]. All studies included participants from both urban and rural locations.

### Risk of Bias

Critical appraisal of the included studies is presented in Table 5. All studies were determined to have a low risk of bias, except for one study that was suspected of having non-response bias [27]. All but one study [31] had problematic reporting of the numerator and denominator, however, this was able to be addressed by the research team by interrogating the provided data or checking source documents from the primary cohort studies. One study was identified as not having an acceptable case definition [21] as it did not specify the period of time covering naturopathy use (e.g. previous 12 months or users’ lifetime).

**TABLE 5:** ASSESSMENT OF RISK OF BIAS AND REPORTING QUALITY FOR INCLUDED STUDIES.

| Criteria | Manuscript |  |  |  |  |  |  |  |
| --- | --- | --- | --- | --- | --- | --- | --- | --- |
|  | Hunt et al (2010) | Klein et al (2015) | Shmueli et al (2010) | Esmail (2017) | Su and Li (2011) | Clarke et al (2015) | McIntyre et al (2019) | Srinivasan and Raji Sugumar (2017) |
| ***Risk of Bias*** |  |  |  |  |  |  |  |  |
| 1 – representativeness of target population | Y | Y | Y | Y | Y | Y | Y | Y |
| 2 – representativeness of sample population | Y | Y | Y | Y | Y | Y | Y | Y |
| 3 – random selection or census | Y | Y | Y | Y | Y | Y | N | Y |
| 4 – non-response bias minimal | Y | Y | N | Y | Y | Y | N | Y |
| 5 – data direct from participants | Y | Y | Y | Y | Y | Y | Y | Y |
| 6 – acceptable case definition | Y | Y | Y | Y | Y | Y | Y | N |
| 7 – reliability and validity of instrument | N/A | N/A | N/A | N/A | N/A | N/A | N/A | N/A |
| 8 – same mode of data for all subjects | Y | Y | Y | Y | Y | Y | Y | Y |
| 9 – appropriate length of shortest prevalence period | Y | Y | Y | Y | Y | Y | Y | N |
| 10 – numerator and denominator appropriate | N | N | N | N | N | N | Y | Y |
| 11 - Summary | Low | Low | Low | Low | Low | Low | Low | Low |
| ***Reporting Quality*** |  |  |  |  |  |  |  |  |
| Title and abstract |  |  |  |  |  |  |  |  |
| 1a – Title | Y | Y | N | N | N | N | N | Y |
| 1b - Abstract | Y | Y | Y | Y | N | N | Y | N |
| Introduction |  |  |  |  |  |  |  |  |
| 2 - Background/rationale | Y | Y | Y | Y | Y | Y | Y | Y |
| 3 - Objectives | Y | Y | Y | Y | Y | Y | Y | Y |
| Methods |  |  |  |  |  |  |  |  |
| 4 - Study design | Y | Y | Y | Y | Y | Y | Y | Y |
| 5 - Setting | Y | Y | Y | Y | Y | Y | Y | Y |
| 6 - Participants | Y | Y | Y | Y | Y | Y | Y | Y |
| 7 - Variables | Y | Y | Y | N | N | Y | Y | N |
| 8 - Data sources/measurement | Y | Y | Y | N | Y | Y | Y | Y |
| 9 - Bias | Y | Y | Y | Y | Y | Y | Y | N |
| 10 - Study size | Y | Y | Y | Y | N | N | Y | Y |
| 11 - Quantitative variables | Y | Y | Y | N | N | Y | Y | N |
| 12a – All statistical methods | Y | Y | N | N | Y | Y | Y | N |
| 12b – Subgroups and interactions | N/A | N/A | N/A | Y | Y | Y | Y | Y |
| 12c – Missing data | N | Y | N | N | N | N | N | N |
| *12d – Analysis accounting for sampling* | N/A | N/A | Y | N | Y | Y | Y | N |
| *12e – Any sensitivity analysis* | N/A | N/A | N/A | N/A | N/A | N/A | N/A | N/A |
| Results |  |  |  |  |  |  |  |  |
| *13a – Numbers of participants* | Y | Y | Y | Y | N | N | Y | N |
| *13b – Reasons for nonparticipation* | N | N | N | N | N | N | N | N |
| *13c – flow diagram* | N | N | N | N | N | N | N | N |
| *14a – Characteristics of study participants* | Y | Y | N | Y | N | Y | Y | Y |
| *14b – Participants with missing data* | N | N | N | N | N | N | N | N |
| *15 - Outcome data* | N | Y | Y | Y | Y | Y | Y | Y |
| *16a – Unadjusted and applicable adjusted estimates* | Y | Y | Y | Y | Y | Y | Y | Y |
| *16b – Report category boundaries* | ? | Y | N/A | N | N/A | N/A | Y | N/A |
| *16c –Estimates of absolute risk* | N/A | N/A | N/A | N/A | N/A | N/A | N/A | N/A |
| *17 - Other analyses* | N/A | N/A | N/A | Y | Y | Y | Y | Y |
| Discussion |  |  |  |  |  |  |  |  |
| *18 - Key results* | Y | Y | Y | Y | Y | Y | Y | N |
| *19 - Limitations* | Y | Y | Y | N | N | N | Y | N |
| *20 - Interpretation* | Y | Y | Y | N | Y | Y | Y | N |
| *21 - Generalisability* | Y | Y | Y | Y | Y | Y | Y | N |
| Other information |  |  |  |  |  |  |  |  |
| *22 - Funding* | Y | Y | Y | Y | N | N | Y | Y |

Assessent of the reporting quality of included studies identified several issues. More than one-half of studies did not clearly identify the study design in the title [21, 27-31]. None of the included studies provided reasons for non-participation or provided information about missing data. Four of the included studies did not acknowledge the limitations of their research. In one case, some of the omissions in reporting may be explained by the nature of the publication (i.e. grey-literature report rather than a peer-reviewed journal article) [30].

### Summary of Findings

The 12-month prevalence reported in studies from the European region ranged between 2% in the UK [25] to 7.7% in Switzerland [26]. One study from the Eastern Mediterranean region (i.e. Israel) [27] reported multiple prevalence rates ranging from 20% in 1993 through to 18% in 2007. Three studies from the Region of the Americas reported 12-month prevalence rates of naturopathy use between 3% (in 1997) and 5% (in 2016) in Canada [30], and between 0.25% (in 2002) and 0.4% (in 2015) in the United States [28, 29]. One study from the Western Pacific region (i.e. Australia) reported a 6.2% prevalence rate [31].

Two studies reported prevalence of naturopathy use over other time periods. One study from the Region of the Americas (Canada) indicated 6% of the general population in 1997, 9% in 2006, and 11% in 2016 used naturopathy at some point in the user’s lifetime [30]. A study from the South-East Asian world region indicated 10% of the population had used naturopathy and yoga, but the timeframe of use was not specified [21].

### Meta-analysis results

The estimated 12-month prevalence rates of naturopathy use for different countries are shown in Figure 2. Prevalence rates significantly differed between countries (p<0.001) and ranged from less than 1% of the population in the USA to 8% in Switzerland. While the primary studies were subject to wide heterogeneity, significant heterogeneity was only found for Canada (p=0.01) and the USA (p<0.001).

**Figure 2:**
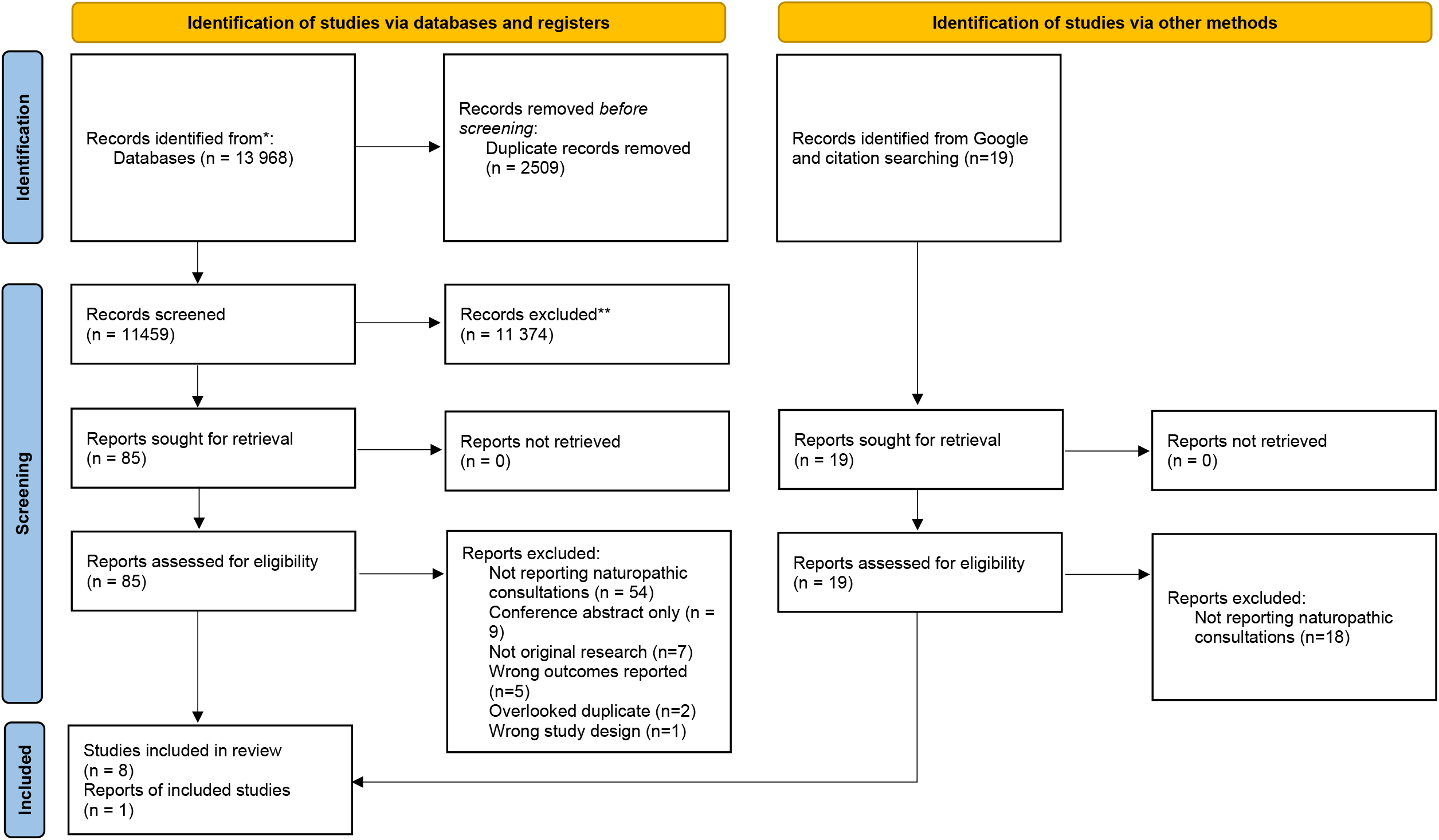
12-month prevalence of naturopathy use in different countries.

Regarding WHO world regions, 12-month prevalence of naturopathy use ranged from 1% in the Region of the Americas to 6% in European and Western Pacific Regions, again with significant differences between regions (p<0.001; Figure 3). Relevant and statistically significant heterogeneity was present in studies involving the European Region (p<0.001), and Region of the Americas (p<0.001).

**Figure 3:**
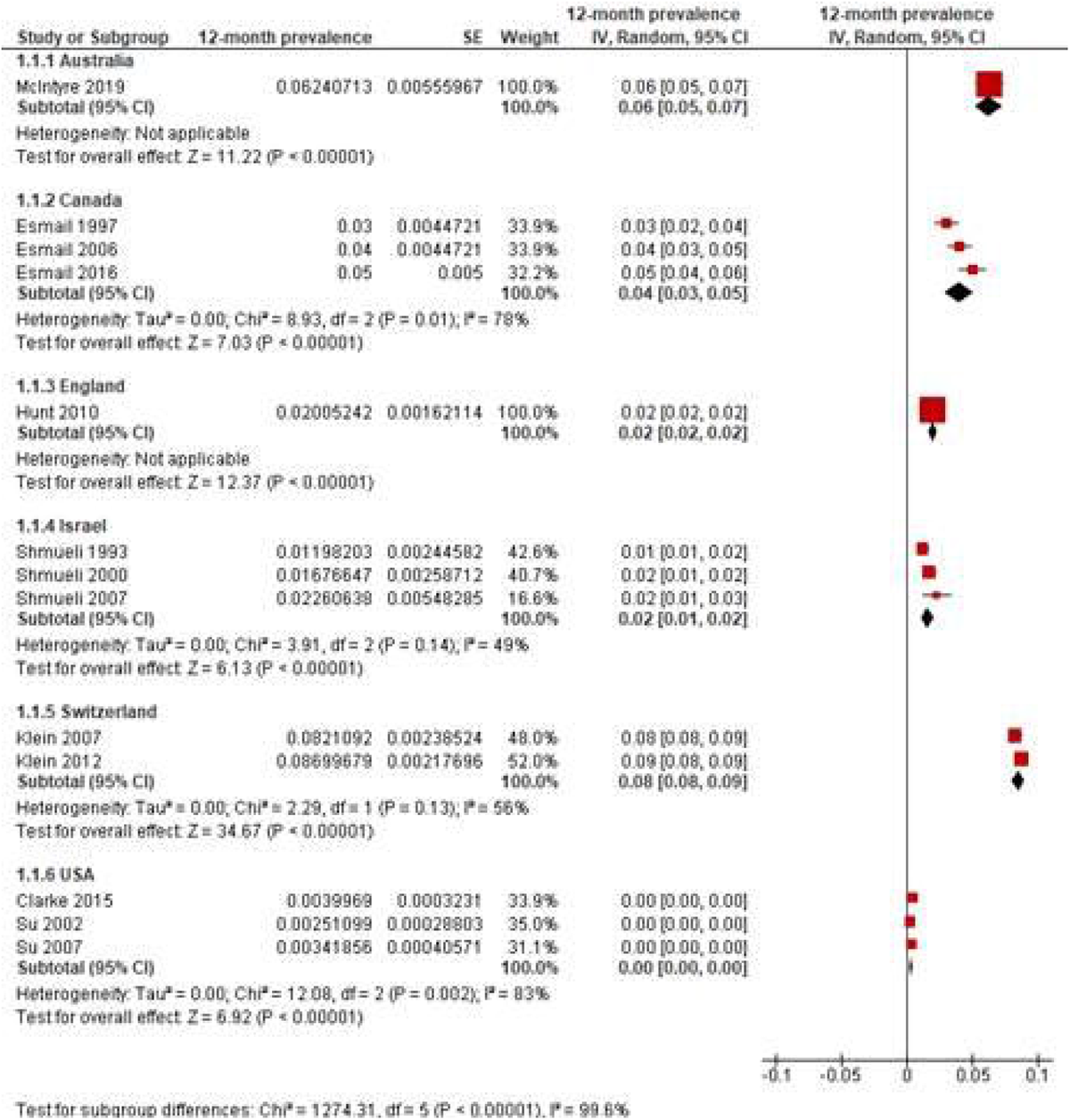
12-month prevalence of naturopathy use in different WHO world regions.

Since all studies were classified as having low risk of bias, no sensitivity analyses were conducted. No meta-analysis could be perfomed on studies reporting prevalence of naturopathy use over other time periods due to the paucity and heterogeneity ofstudies reporting this outcome.

## Discussion

This review presents the most recent synthesis of evidence of the global prevalence of consultations with naturopaths/naturopathic doctors. The prevalence of naturopathy/naturopathic medicine use was reported in seven countries, across five WHO designated regions of the world. Of the regions reporting 12-month prevalence rates, the highest was in the Eastern Mediterranean region (Israel), with 18% (2007) to 20% (1993) of the general population seeking the services of a naturopath/naturopathic doctor. The lowest reported 12-month prevalence of naturopathy use was observed in the Americas (USA), with a rate of 0.4% (2012). Lifetime prevalence of use was reported in two countries: Canada (6% in 1997 to 11% in 2016); and India (7% rural, 12% urban in 2011/12). Where more than one timeframe of data was available, there was a relative amount of consistency across time suggesting naturopathy/naturopathic medicine use is temporally stable in these countries.

The wide range in the rates of consultation with a naturopath/naturopathic doctor may reflect differences in the perception and availability of naturopathy in specific countries. For example, while national prevalence of consultations with naturopaths in the USA is relatively low, this may obscure significant heterogeneity within that region. For example, insurance data from Washington state shows prevalence of naturopathic consultation to be four times higher than the national prevalence (1.6% v 0.4%) [32]. Such heterogeneity may be similarly observed in other regions and may be due to several factors. In the USA recognition of the naturopathic profession through licensure is not uniformly applied across that nation [33], and distribution of the naturopathic workforce has historically been determined by the proximity to naturopathic educational institutions [34]. Insurance coverage is also known to be a significant driver of naturopathic use [32], and variable insurance coverage arrangements for naturopathy – as observed in the USA [35] – may also result in regional differences. Further attention towards regional variations and heterogeneity, particularly as it relates to specific barriers and facilitators to appropriate utilization of naturopathic services - is warranted.

The wide range in rates of naturopathy use may also reflect differences in scope of practice in each world region. For example, in the USA, naturopathic physicians are considered to bridge conventional medicine and CAM modalities [36], while in Germany, naturopathic practitioners known as “Heilpraktiker” are a distinct category and reportedly have inconsistent training and clinical abilities [37]. As such, the term naturopathy may be differentially classifying practitioners due to professionalization, resulting in an underestimate of use in some countries and overestimate in others. Further consideration of the implications associated with the inconsistent ‘protection’ of professional titles and defined scopes of practice for naturopaths/naturopathic doctors by country is likely to influence the prevalence of use by the public [2].

Prevalence data from some countries may also be impacted by definitional difficulties or confusion around the term ‘naturopathy’. For example, naturopathy is often grouped under a broader nomenclature as one of the many modalities or therapies considered ‘complementary approaches to healthcare’ [38] or “integrative medicine” and thus may not be individually represented in the publications included in our analysis. Multiple practitioner types may also present difficulties for data collection. For example, a review of CAM services in Europe, of the (22,300) practitioners of naturopathy, 15,000 were identified as (mostly German) medical doctors [39]. Thus, patients may not identify obtaining naturopathy as a service per se, but as part of the standard care they receive from a medical doctor who integrates naturopathic principles or modalities into their practice. This may be one reason why three of the largest European countries by naturopathic workforce (Germany, Portugal and Spain [2]) were not represented in this review. Thus, the true prevalence of naturopathic consultations is likely under-reported. Further, an examination of government administered national health surveys of the general population in the countries represented by WNF member organisations, found only Switzerland, Northern Ireland, USA, Mexico and India currently included items that specifically measured consultations with a naturopath/naturopathic doctor (see Supplementary File 2). To evaluate the potential role of naturopaths in care delivery, it is imperative that naturopathic health services and workforce research data is captured in all countries where there is a significant naturopathic presence.

Furthermore, although naturopathic practice is relatively consistent globally, local, and regional variations in preferred therapies may result in point-of-service differences that may impact prevalence of naturopathic consultations in those countries. For example, in the United Kingdom, historical connections between osteopathy and naturopathy may drive naturopathic use for musculoskeletal conditions in that country more than in countries like Australia, where naturopathy and herbalism have had a larger shared history and connection [40]. Some studies in this review explicitly combined queries about naturopathic utilization with other CAM practices – for example, herbalism and naturopathy in the Australian study. Thus, it is important that a reliable validated instrument is developed for collecting more specific data about naturopathic service utilization within and across countries to establish ‘true’ prevalence of use information.

While prevalence data provides a snapshot of a given populations’ use of naturopathy, less is known about the factors associated with that use. For example, factors that have previously been raised as impacting the use of naturopathy/naturopathic medicine, include licensure and regulation, scope of practice, training of new students and therefore number of naturopaths/naturopathic doctors in the workforce, or country specific health systems that influence the support and reimbursements of naturopathic services (e.g. insurance vs out of pocket) [41]. By focusing on general population utilization, this study may also not reflect differences in prevalence of use for different clinical conditions. For example, Australian studies published before 2010 show a self-reported prevalence of naturopathic use among the general population of mid-aged women to be 8.7%, while rates for cancer (15.7%) and depression (22.2%) were significantly higher [9]. Similar variations were seen in insurance data from Washington state in the US, where 7.1% of insured cancer patients made claims for naturopathic treatment, compared to 1.6% of general enrollees [32].

One of the limitations of prevalence studies in the context of naturopathy, is they fail to capture the breadth of treatments that is unique to naturopathy and they do not capture data associated with the quality of care, role within healthcare systems, nor the efficacy and safety of naturopathic approaches to the management of specific conditions [42]. Thus, research into the quality, safety, efficacy, and cost effectiveness of naturopathy/naturopathic medicine would provide pragmatic understanding about the contribution of naturopathy to healthcare within populations and more broadly across the world. Additionally, although limiting data collection to studies published after 2010 helps to ensure prevalence data most accurately reflects contemporary utilization, such time limits may have excluded some studies in regions that were missing from the review. Additionally, observing changes in prevalence of naturopathic consultations over time may also be able to offer insights into the changing role of naturopathy/naturopathic medicine in relation to health systems changes or generational health needs [43].

## Conclusion

Although the naturopathic workforce has a significant presence globally, there is limited detailed data on the prevalence of naturopathic consultations. As such, there is a need for a reliable validated instrument to be developed for collecting more specific data about naturopathic service utilization within and across countries. Nevertheless, current evidence reports a 12-month prevalence of naturopathy use ranging from 1% in the Region of the Americas to 6% in European and Western Pacific Regions, though there are significant differences between and within world regions. Differences in naturopathic utilization in these regions may be indicative of a range of policy, legislative and social factors impacting the naturopathic profession. Despite these ongoing factors, further research attention is warranted to support the integration of naturopathic services into healthcare systems to ensure consumers have access to safe and effective multi-disciplinary care.

## Data Availability

All data used in the analysis presented in this paper is publicly available and can also be accessed by contacting the authorship team.

## Author contributions

AS devised the project, the main conceptual idea and drafted the review protocol. All authors reviewed and edited the protocol prior to registration. Literature searching, removal of duplicates and filtering of citations by title and abstract was undertaken by AM. Full text retrieval and assessment of articles against eligibility criteria was undertaken by AM and AS. Data extraction was completed by JHar. STROBE assessment was completed by CVV, JS and AS. Risk of bias assessment was completed by JG, JHaw and AS. Meta-analysis was completed by JG, KC and HC. The method section of the manuscript was drafted by HC, AS, JS and JHaw. The results were drafted by AS and HC. The discussion was drafted by JHar, JW, JA and BL. The introduction was drafted by RR, ML, and RB. All authors reviewed and edited the full draft of the manuscript prior to submission.

## Competing interests

The authors have no competing interests to declare.

## Funding

This research received no specific grant from any funding agency in the public, commercial or not-for-profit sectors.

## Notes

### Competing Interest Statement

The authors have declared no competing interest.

### Author Declarations

UTS Human Research Ethics Committee

## References

1. Snider, P. and J. Zeff, Unifying Principles of Naturopathic Medicine Origins and Definitions. Integrative Medicine: A Clinician’s Journal, 2019. 18(4): p. 36.

2. World Naturopathic Federation Roots Committee, WNF - Naturopathic Roots Report. 2016, World Naturopathic Federation,: Toronto, Canada.

3. Dunn, J., et al., Characteristics of global naturopathic education, regulation, and practice frameworks: results from an international survey. BMC complementary medicine and therapies, 2021. 21(1): p. 1–19.

4. Steel, A., et al., Overview of international naturopathic practice and patient characteristics: results from a cross-sectional study in 14 countries. BMC Complementary Medicine and Therapies, 2020. 20(1): p. 59.

5. World Naturopathic Federation, 2016 Naturopathic Numbers Report. 2016, World Naturopathic Federation: Canada.

6. World Naturopathic Federation, Global Naturopathic Regulation. 2018, World Naturopathic Federation: Toronto, Ontario.

7. Smith, M., A. Burton, and T. Falkenberg, World Health Organization Traditional Medicine Strategy 2014–2023. New strategy for traditional and complementary medicine includes the development and use of herbal medicinal preparations. HerbalEGram, 2014. 11(5).

8. Leach, M.J., Profile of the complementary and alternative medicine workforce across Australia, New Zealand, Canada, United States and United Kingdom. Complementary therapies in medicine, 2013. 21(4): p. 364–378.

9. Reid, R., et al., Complementary medicine use by the Australian population: a critical mixed studies systematic review of utilisation, perceptions and factors associated with use. BMC Complementary and Alternative Medicine, 2016. 16(1): p. 176.

10. Leach, M.J., Determinants of Complementary Medicine Service Utilization in a Regional South Australian Population: A Cross-Sectional Study. The Journal of Rural Health, 2021. 37(1): p. 69–80.

11. Bradley, R., et al., Survey of CAM interest, self-care, and satisfaction with health care for type 2 diabetes at group health cooperative. BMC complementary and alternative medicine, 2011. 11(1): p. 121.

12. Foley, H. and A. Steel, Patient perceptions of clinical care in complementary medicine: A systematic review of the consultation experience. Patient Educ Couns, 2016. 100: p. 212–23.

13. Foley, H., A. Steel, and J. Adams, Perceptions of Person-Centred Care Amongst Individuals with Chronic Conditions who Consult Complementary Medicine Practitioners. Complementary Therapies in Medicine, 2020: p. 102518.

14. Foley, H., A. Steel, and J. Adams, Consultation with complementary medicine practitioners by individuals with chronic conditions: Characteristics and reasons for consultation in Australian clinical settings. Health & social care in the community, 2020.

15. Legenne, M., et al., Perception of naturopathy for female patients with metastatic gynecological cancer: A qualitative study. Palliative & supportive care, 2015. 13(6): p. 1663–1668.

16. Shea, B.J., et al., AMSTAR 2: a critical appraisal tool for systematic reviews that include randomised or non-randomised studies of healthcare interventions, or both. bmj, 2017. 358.

17. Innovation, V.H., Covidence systematic review software. Melbourne, Australia.

18. Knottnerus, A. and P. Tugwell, STROBE--a checklist to Strengthen the Reporting of Observational Studies in Epidemiology. Journal of clinical epidemiology, 2008. 61(4): p. 323.

19. Hsu, C., et al., New perspectives on patient expectations of treatment outcomes: results from qualitative interviews with patients seeking complementary and alternative medicine treatments for chronic low back pain. BMC Complementary Alternative Medicine, 2014. 14(1): p. 276.

20. Organization, W.H., WHO global report on traditional and complementary medicine 2019, in WHO global report on traditional and complementary medicine 2019. 2019.

21. Srinivasan, R. and V.R. Sugumar, Spread of traditional medicines in India: Results of national sample survey organization’s perception survey on use of Ayush. Journal of evidence-based complementary & alternative medicine, 2017. 22(2): p. 194–204.

22. Neyeloff, J.L., S.C. Fuchs, and L.B. Moreira, Meta-analyses and Forest plots using a microsoft excel spreadsheet: step-by-step guide focusing on descriptive data analysis. BMC research notes, 2012. 5(1): p. 1–6.

23. Higgins, J.P., et al., Measuring inconsistency in meta-analyses. Bmj, 2003. 327(7414): p. 557–560.

24. Higgins, J.P., et al., Cochrane handbook for systematic reviews of interventions. 2019: John Wiley & Sons.

25. Hunt, K.J., et al., Complementary and alternative medicine use in England: Results from a national survey. International Journal of Clinical Practice, 2010. 64(11): p. 1496–1502.

26. Klein, S.D., et al., Usage of complementary medicine in Switzerland: Results of the Swiss health survey 2012 and development since 2007. PLoS ONE, 2015. 10(10): p. no pagination.

27. Shmueli, A., I. Igudin, and J. Shuval, Change and stability: use of complementary and alternative medicine in Israel: 1993, 2000 and 2007. European Journal of Public Health, 2011. 21(2): p. 254–259.

28. Su, D. and L. Li, Trends in the use of complementary and alternative medicine in the United States: 2002-2007. Journal of Health Care for the Poor and Underserved, 2011. 22(1): p. 296–310.

29. Clarke, T.C., et al., Trends in the use of complementary health approaches among adults: United States, 2002-2012. National health statistics reports, 2015(79): p. 1–16.

30. Esmail, N., Complementary and Alternative Medicine. 2017: Fraser Institute.

31. McIntyre, E., et al., Consultations with Naturopaths and Western Herbalists: Prevalence of Use and Characteristics of Users in Australia. The Journal of Alternative Complementary Medicine, 2019. 25(2).

32. Lafferty, W.E., et al., The use of complementary and alternative medical providers by insured cancer patients in Washington State. Cancer, 2004. 100(7): p. 1522–30.

33. Fleming, S.A. and N.C. Gutknecht, Naturopathy and the primary care practice. Primary Care: Clinics in Office Practice, 2010. 37(1): p. 119–136.

34. Albert, D.P. and F.B. Butar, Distribution, concentration, and health care implications of naturopathic physicians in the United States. Complementary health practice review, 2004. 9(2): p. 103–117.

35. Whedon, J., et al., Insurance reimbursement for complementary healthcare services. The Journal of Alternative and Complementary Medicine, 2017. 23(4): p. 264–267.

36. Litchy, A.P., Naturopathic physicians: holistic primary care and integrative medicine specialists. Journal of dietary supplements, 2011. 8(4): p. 369–377.

37. Heudorf, U., A. Carstens, and M. Exner, Naturopathic practitioners and the public health system. Legal principles as well as experience from naturopathic practitioner candidate tests and hygiene inspections of naturopathic practitioner’s practices in the Rhine-Main area in 2004-2007. Bundesgesundheitsblatt, Gesundheitsforschung, Gesundheitsschutz, 2010. 53(2): p. 245–257.

38. National Center for Complementary and Integrative Health. Are You Considering a Complementary Health Approach? 2016 [cited 2021 30 June]; Available from: https://www.nccih.nih.gov/health/are-you-considering-a-complementary-health-approach.

39. von Ammon, K., et al., Complementary and alternative medicine provision in Europe–first results approaching reality in an unclear field of practices. Complementary Medicine Research, 2012. 19(Suppl. 2): p. 37–43.

40. World Naturopathic Federation, World Naturopathic Federation Report. 2015, World Naturopathic Federation: Ontario, Canada.

41. Ooi, S.L., L. McLean, and S.C. Pak, Naturopathy in Australia: Where are we now? Where are we heading? Complementary therapies in clinical practice, 2018. 33: p. 27–35.

42. Myers, S. and V. Vigar, The State of the Evidence for Whole-System Multi-Modality Naturopathic Medicine: A Systematic Scoping Review. The Journal of Alternative & Complementary Medicine, 2019. 25(2).

43. Steel, A., et al., Generational differences in complementary medicine use in young Australian women: Repeated cross-sectional dataset analysis from the Australian longitudinal study on women’s health. Complementary therapies in medicine, 2019. 43: p. 66–72.

