## Supplementary File 1 for "INTERNATIONAL PREVALENCE OF CONSULTATION WITH A NATUROPATHIC PRACTITIONER: A SYSTEMATIC REVIEW AND META-ANALYSIS"

*Supplementary File 2: List of national surveys from WNF member countries, with reference to inclusion of items examining naturopathy use*

| Country | Report/survey identified/ located | Inclusion of naturopathy-specific item | Prevalence timeframe | Date last collected | Other dates collected | Item/s | Data accessibility |
|---|---|---|---|---|---|---|---|
| **FULL MEMBERS** | | | | | | | |
| Australia | National Health Survey | Absent | | 2021 | | | |
| Belgium | Health Interview Survey https://his.wiv-isp.be/fr/Documents%20partages/Summ_HC_FR_2018.pdf | Absent | | 2018 | Every 2 years from 1997 | | |
| Brazil | National Health Survey - PNS Table 3.21 https://www.ibge.gov.br/en/statistics/social/health/16840-national-survey-of-health.html?=&t=downloads | Absent | | 2019 | 2013 | | Appears to be available at link https://www.ibge.gov.br/en/statistics/social/health/16840-national-survey-of-health.html?=&t=downloads |
| Canada | Canadian Health Measures Survey | Absent | | 2019 | Every 2 years since 2011 | - | |
| Canada | Canadian National Health Survey | Absent | - | 2016 | | - | CNHS: https://www.statcan.gc.ca/eng/surveys?MM=1 |

| | | | | | | | |
|---|---|---|---|---|---|---|---|
| Canada | National Population Health Survey | Present but combined with other health service (homeopathy ) | 12 month use | 2010/11 | Every 2 years since 1992 | A) People may also use alternative or complementary medicine. In the past 12 months, [have/has] [you/FNAME] seen or talked on the telephone to an alternative health care provider such as an acupuncturist, homeopath or massage therapist about [your/his/her] physical, emotional or mental health?<br>B) Who did you speak to (answer option is "homeopath or naturopath" | https://crdcn.org/datasets/nphs-national-population-health-survey<br>https://crdcn.org/research<br><br>Application process for academic researchers<br>Researchers wishing to access the RDC should create an account on the Statistics Canada Microdata Access Platform and follow the steps to create a new proposal. The proposal is evaluated by Statistics Canada for feasibility before access can be granted. In addition, if you are a student, your thesis supervisor must write a letter in support of your RDC application and join the application as a co-investigator. For other academic users, a completed peer-review may be required. The review must be conducted by a tenured faculty-member at an accredited Canadian university. Researchers who are required to submit such a peer review can source their own peer reviewer, or contact CRDCN for assistance if they are unable to find a suitable candidate.<br>Access fees for certains users<br>Fees can apply to certain research projects conducted in the RDCs. Consult the Access & Fee-For-Service Policy to learn more. |
| Chile | National Health Survey | Absent | | Unclear - maybe 2016/17 | Every 4 years | Appears to exist, but cannot locate a recent copy of the survey or results. An earlier version (2009-10) suggests use of CAM was assessed, but all CAM were grouped together as one variable. https://www.who.int/fctc/reporting/party_reports/chile_annex1_national_health_survey_2010.pdf | Maybe somewhere on this site (might need a Spanish-speaker):<br>https://deis.minsal.cl/#estadisticas |
| Cyprus | "State of health" survey | Absent | | 2019 | | | https://ec.europa.eu/health/sites/health/files/state/docs/2019_chp_cyprus_english.pdf<br>https://www.euro.who.int/__data/assets/pdf_file/0007/355975/Health-Profile-Cyprus-Eng.pdf |

| | | | | | | | |
|---|---|---|---|---|---|---|---|
| Democratic Republic of Congo | DHS Demographic and Health Survey | Absent | | 2013-2014 | | | https://www.dhsprogram.com/pubs/pdf/FR300/FR300.pdf |
| Ecuador | Health survey | Absent | | 2017 | | | https://www.ecuadorencifras.gob.ec/documentos/web-inec/Estadisticas_Sociales/Recursos_Actividades_de_Salud/RAS_2017/Principales_Resultados_%28RAS%29.pdf |
| Egypt | DHS Demographic and Health survey | Absent | | 2015 | 2014, 2008, 2005 | | https://dhsprogram.com/pubs/pdf/FR313/FR313.pdf |
| El Salvador | National Family Health Survey | Absent | | 2008 | | | file:///C:/Users/User/AppData/Local/Temp/Cuestionario_El%20Salvador%202008_Nombre%20de%20variables.pdf |
| France | National Health and Nutrition Survey | Absent | | 2006 | | | file:///C:/Users/User/AppData/Local/Temp/26327_7069-rapp-inst-enns-web.pdf |
| Greece | Hellenic National Nutrition and Health Survey | Absent | | 2013-2015 | | | |
| Greece | World Health Survey | Absent | | 2003 | | | |
| Greece | Greek National Survey on Health and Nutrition (the HYDRIA Proejct) | Absent | | 2009-2011 | | | |
| Hong Kong | Population Health Survey and Health Behaviour Survey | Absent | | 2018/19 (report not yet released) | 2014/15, 2003/04 | | |
| India | DHS Demographic and Health survey | Absent | | 2019-20 | 2015-16, 2005-06, 1998-99, 1992-93 | | |

| | | | | | | | |
|---|---|---|---|---|---|---|---|
| India | NFHS - National Family Health Survey | Present but combined with other health service (yoga) | Generally used when sick (household questionnaire)<br><br>Men's and women's questionnaires also asks about places to receive family planning, where they take children when sick, and a number of other specific details relating to health care utilisation around family planning. | 2019-20 | 2015-16<br>2005-06<br>1998-99<br>1992-93 | Q. When members of your household get sick, where do they generally go for treatment?<br>A. (option) Yoga and Naturopathy<br>[also separates into public and private] | Process is unclear?<br>http://rchiips.org/nfhs/data1.shtml |
| India | AHS - Annual Health Survey | Absent | | | | | |
| Italy | ISSP - International Social Survey Programme: Health and Health Care | Absent | 12 month use | 2011 | | During the past 12 months, how often did you visit or were visited by... an [alternative/traditional /folk]health care practitioner? | https://search.gesis.org/research_data/ZA5800 |
| Italy | Italy National Healthy Survey | **Unknown due to survey availability** | | | | | |

| | | | | | | | |
|---|---|---|---|---|---|---|---|
| Italy | EHIS - European Health Interview Survey | Absent | | 2019 | 2015 | | https://www.istat.it/en/archivio/210553 |
| Japan | The Japan National Health and Nutrition Survey (NHNS) | Absent | | | | | |
| Malaysia | National Health and Morbidity Survey | Absent | | | | | https://iptk.moh.gov.my/images/technical_report/2020/FactSheet_BI_AUG2020.pdf |
| Mexico | National Health Survey (ENSA) | Present (as 'Naturista') | Unclear | 2018-19 | 2016<br>2012<br>2006 | Q4.8: https://en.www.inegi.org.mx/contenidos/programas/ensanut/2018/doc/ensanut_2018_cuestionario_hogar.pdf | https://en.www.inegi.org.mx/programas/ensanut/2018/ |
| Nepal | DHS Demographic and Health survey | Absent | | | | | |
| Nepal | Noncommunicable Disease Risk Factors: STEPS Survey Nepal 2019 | Present but combined with other health services (traditional medicine) | For specific health conditions - Normal source of treatment<br>For smoking cessation - 12 month use | 2019 | | "During the past 12 months, what did you do to try and stop smoking?"<br>"Where do you usually go for treatment or advice for you >condition<?"<br>"Where do you usually get your drugs for >condition<?" | https://www.who.int/docs/default-source/nepal-documents/ncds/ncd-steps-survey-2019-compressed.pdf?sfvrsn=807bc4c6_2 |
| New Zealand | New Zealand Health Survey | Absent | | | | | https://www.health.govt.nz/publication/questionnaires-and-content-guide-2019-20-new-zealand-health-survey |
| Nigeria | DHS Demographic and Health survey | Absent | | | | | |
| Peru | ENCUESTA DEMOGRÁFICA Y DE SALUD FAMILIAR (ENDES) | Absent | | | | | http://iinei.inei.gob.pe/microdatos/ |
| Portugal | National Health Survey | Absent | | 2019 | 2018<br>2017<br>2016<br>etc. annually | | https://www.ine.pt/xportal/xmain?PORTLET_ID=JSP&xpgid=ine_publicacoes&xpid=INE&PORTLET_NAME=ine_cont_header_pub_en&PORTLET_UID=%23JSP%3Aine_cont_header_pub_en%23&PUBLICACOEStema=00&PUBLICACOESdata_inicial=01-07-2014&PUBLICACOESdata_final=13-07-2021&x=14&y=10&PUBLICACOESfreeText=health |

| | | | | | | | |
|---|---|---|---|---|---|---|---|
| Puerto Rico | Unknown | | | | | | |
| Russia | Longitudinal Monitoring Survey of HSE (health service questions in Adult survey) | Absent | | 2019 | 1994 onward | | https://rlms-hse.cpc.unc.edu/ |
| Russia | Kantar National Health and Wellness Survey | Unknown due to survey availability | | 2011 | | | https://www.kantar.com/expertise/health/da---real-world-data-pros-claims-and-health-records/national-health-and-wellness-survey-nhws |
| Saudi Arabia | World Health Survey Saudi Arabia (KSAWHS) | Absent | | 2019 | | | https://www.moh.gov.sa/en/Ministry/Statistics/Population-Health-Indicators/Documents/World-Health-Survey-Saudi-Arabia.pdf |
| Saudi Arabia | Saudi Health Interview Survey | Absent | | 2013 | | | http://www.healthdata.org/sites/default/files/files/Projects/KSA/Saudi-Health-Interview-Survey-Results.pdf |
| Saudi Arabia | Saudi Health Interview Census | Unknown due to survey availability | | 2015 | | | |
| Slovenia | World Health Survey | Absent | | | | | |
| Slovenia | European Health Interview Survey | Absent | | 2007 | | | https://www.stat.si/doc/pub/IVZ-angl.pdf https://ec.europa.eu/eurostat/web/microdata/european-health-interview-survey |
| South Africa | South Africa Demographic and Health Survey (DHS) | Absent | | 2016 | 2003 | | https://dhsprogram.com/pubs/pdf/FR337/FR337.pdf |
| South Africa | South African Health and Nutrition Examination Survey (SANHANES-1) | Absent | | 2012 | | | file:///C:/Users/User/AppData/Local/Temp/7844.pdf |
| Spain | National Health Survey | Absent | | 2017 | 2011-12<br>2006<br>2003 | | |

| | | | | | | | |
|---|---|---|---|---|---|---|---|
| Switzerland | Swiss Health Survey | **Present** | 12 month use | 2017 | 2012<br>2007 | How often have you been to one of the following specialists in the last 12 months: Naturopath | Available from the Swiss Federal Statistical Office http://www.bfs.admin.ch/bfs/portal/de/index/infothek/erhebungen__quellen/blank/blank/ess/04.html<br><br>2012 and 2007 data reported here: https://journals.plos.org/plosone/article?id=10.1371/journal.pone.0141985 |
| United Kingdom - England | Health Survey for England (HSE) | Absent | | 2019 | Annually | | https://digital.nhs.uk/data-and-information/publications/statistical/health-survey-for-england/2019 |
| United Kingdom - Scotland | Scottish Health Survey | Absent | By health condition, 12 month use | 2020 | Annually | Have you received any treatment advice for >insert condition< from any of the people on this card: Other alternative medicine professional | https://beta.ukdataservice.ac.uk/datacatalogue/studies/study?id=8737#!/documentation |
| United Kingdom - Wales | National Survey for Wales | Absent | By health condition, 12 month use<br><br>Non-GP primary care, 12 month use | Rolling (monthly interviews) | | In the last 12 months, which of these kinds of treatment or management have you had for >insert condition<: Complementary therapies (e.g. acupuncture, massage)<br><br>In the last 12 months, which of these services have you used for yourself: Osteopath | https://gov.wales/national-survey-wales-questionnaires |
| United Kingdom - Northern Ireland | Health Survey Northern Ireland | Absent | | 2019-20 | Annually | | https://www.data-archive.ac.uk/home |

| | | | | | | | |
|---|---|---|---|---|---|---|---|
| United Kingdom - Northern Ireland | Northern Ireland Life and Times Survey (I don't think this is actually a government survey - run by Queen's University Belfast and Ulster University) | **Present** - but only in 2005 | Use ever | 2005 | Annually, but CAM only covered in 2005 | Have you ever used naturpathy? | https://www.ark.ac.uk/nilt/datasets/ https://www.ark.ac.uk/nilt/2005/Complementary_Medicine/COMTH8.html |
| Uruguay | Uruguay Continuous Household Survey | Absent | | 2020 | Annually | | https://www.ine.gub.uy/encuesta-continua-de-hogares1 |
| USA | National Health Interview Survey - CAM Supplement | **Present** | 12 month use | 2012 | 2007 2002 | | https://www.cdc.gov/nchs/nhis/data-questionnaires-documentation.htm https://www.ncbi.nlm.nih.gov/pmc/articles/PMC4573565/ |
| Zambia | DHS Demographic and Health Survey | Absent | | 2018-19 | | | https://microdata.worldbank.org/index.php/catalog/3597 |
| **ASSOCIATE MEMBERS** | | | | | | | |
| Ireland | SLÁN - Survey of Lifestyle, Attitudes and Nutrition | Absent | Use ever and 12 month use | 2007 | 2002 1998 | Have you ever attended an alternative/complementary practitioner? (e.g. acupuncturist, homeopath, reflexologist) | https://www.ucd.ie/issda/data/surveyonlifestyleandattitudestonutritionslan/ |
| Ireland | Healthy Ireland | Absent | | 2018 | 2017 2016 2015 | | https://www.ucd.ie/issda/data/healthyireland/ |

| | | | | | | | |
|---|---|---|---|---|---|---|---|
| Norway | HUNT - The Trondelag Health Study<br><br>(Norway also has research centre - NAFKAM - which conducts national surveys on CAM, but they don't cover naturopathy in their list of professions https://nafkam.no/en/report-use-complementary-and-alternative-medicine-cam-norway-2018 ) | Absent | 12 month use | | | HUNT 2 - During the last 12 months, have you visited any of the following: Other treatment provider (naturopath, reflexologist….)<br><br>HUNT 3, CAM suppl - How many times in the last 12 months have you been to an alternative practitioner?<br>Which type of alternative treatment did you receive and who did you receive the treatment from?: Other type of alternative treatment | https://www.ntnu.edu/hunt/research<br>https://www.ntnu.edu/hunt/data/que |
| Singapore | National Population Health Survey | Absent | | 2018-19 | 2016-17 | | https://www.moh.gov.sg/docs/librariesprovider5/default-document-library/nphs-2019-survey-report.pdf |
| Singapore | National Health Surveillance Survey | Absent | | 2007 | 2001 | | https://www.singstat.gov.sg/find-data/search-by-theme/society/health/latest-data |
| Singapore | Singapore National Health Survey | Absent | | 2010 | 2004<br>1998 | | https://www.singstat.gov.sg/find-data/search-by-theme/society/health/latest-data |
| **EDUCATIONAL MEMBERS** | | | | | | | |
| Czech Republic | HELEN (Health, Lifestyle and Environment) Study | Absent | | 2014 | Annually since 2003 | | http://www.szu.cz/publikace/studie-helen?lang=1 |
| Czech Republic | World Health Survey | Absent | | 2003 | | | https://microdata.worldbank.org/index.php/catalog/1703 |
| Ghana | DHS Demographic and Health Survey | Absent | | 2017 | 2014 | | https://dhsprogram.com/methodology/survey/survey-display-506.cfm |
