## Supplementary File 2 for "INTERNATIONAL PREVALENCE OF CONSULTATION WITH A NATUROPATHIC PRACTITIONER: A SYSTEMATIC REVIEW AND META-ANALYSIS"

*Supplementary File 1: List of Excluded articles and reasons for exclusion*

| Title | Authors | Published Year | Journal | Volume | Issue | Pages | Notes | Tags |
|---|---|---|---|---|---|---|---|---|
| **Trends in the use of complementary health approaches among adults: United States, 2002-2012** | Clarke, T. C.; Black, L. I.; Stussman, B. J.; Barnes, P. M.; Nahin, R. L. | 2015 | National health statistics reports | | 79 | Jan-16 | Exclusion reason: Does not report naturopathic consultations | |
| **The prevalence and experience of Australian naturopaths and Western herbalists working within community pharmacies** | | 2011 | Journal of the Australian Traditional-Medicine Society | 17 | 4 | 240-240 | Exclusion reason: Duplicate | |
| **Why seek complementary medicine? An observational study in homeopathic, acupunctural, naturopathic and mainstream medical practice** | Van Dulmen, S.; De Groot, J.; Koster, D.; Heiligers, P. J. M. | 2010 | Journal of Complementary and Integrative Medicine | 7 | 1 | 20 | Exclusion reason: Does not report naturopathic consultations; | |
| **The Australian Complementary Medicine Workforce: A Profile of 1,306 Practitioners from the PRACI Study** | Steel, A.; Leach, M.; Wardle, J.; Sibbritt, D.; Schloss, J.; D. Iezel H; Adams, J. | 2018 | Journal of Alternative and Complementary Medicine | 24 | 4 | 385-394 | Exclusion reason: Does not report naturopathic consultations; | |
| **Primary Care in Oregon: The Naturopathic Physician's Perspective** | Linn, Brooke L.; Metcalf, Gary | 2018 | | | 10979746 | 231 | Exclusion reason: Not original research; | |
| **Characteristics of the Australian complementary and alternative medicine (CAM) workforce** | Leach, Matthew J.; McIntyre, Erica; Frawley, Jane | 2014 | Australian Journal of Herbal Medicine | 26 | 2 | 58-65 | Exclusion reason: Does not report naturopathic consultations; | |
| **[Which complementary and alternative medicine modalities are integrated within Israeli healthcare organizations and do they match the public's preferences?]** | Keshet, Y.; Ben-Arye, E. | 2011 | Harefuah | 150 | 8 | 635-689 | Exclusion reason: Does not report naturopathic consultations; | |
| **Complementary medical health services: a cross sectional descriptive analysis of a Canadian naturopathic teaching clinic** | Kennedy, Deborah A.; Bernhardt, Bob; Snyder, Tara; Bancu, Viviana; Cooley, Kieran | 2015 | BMC Complementary & Alternative Medicine | 15 | 1 | 1-Oct | Exclusion reason: Wrong outcomes; | |
| **Characteristics and job satisfaction of general practitioners using complementary and alternative medicine in Germany--is there a pattern?** | Joos, Stefanie; Musselmann, Berthold; Szecsenyi, Joachim; Goetz, Katja | 2011 | BMC Complementary and Alternative Medicine | 11 | | 131 | Exclusion reason: Wrong outcomes; | |
| **Naturopathic practice at North American academic institutions: Description of 300,483 visits and comparison to conventional primary care** | Chamberlin, S. R.; Oberg, E.; Hanes, D. A.; Calabrese, C. | 2014 | Integrative Medicine Insights | | 9 | Jul-15 | Exclusion reason: Wrong outcomes; | |
| **Complementary and alternative medicine among Filipinos: Prevalence, costs and patterns of use** | Morfe, J. H. D.; Lim, V. S. | 2013 | Phillippine Journal of Internal Medicine | 51 | 4 | | Exclusion reason: Wrong study design; | |

| | | | | | | | |
|---|---|---|---|---|---|---|---|
| **The prevalence and experience of Australian naturopaths and Western herbalists working within community pharmacies** | Braun, L. A.; Spitzer, O.; Tiralongo, E.; Wilkinson, J. M.; Bailey, M.; Poole, S.; Dooley, M. | 2011 | BMC Complementary and Alternative Medicine | 11 | 41 | (23 May 2011) | Exclusion reason: Wrong outcomes; |
| **Integration of complementary and alternative medicine into family practices in Germany: Results of a national survey** | Joos, S.; Musselmann, B.; Szecsenyi, J. | 2011 | Evidence-based Complementary and Alternative Medicine | 2011 | | 495813 | Exclusion reason: Wrong outcomes; |
| **USE OF COMPLEMENTARY AND ALTERNATIVE MEDICINE IN GEORGIA** | Nadareishvili, I.; Lunze, K.; Tabagari, N.; Beraia, A.; Pkhakadze, G. | 2017 | Georgian Medical News | | 272 | 157-164 | Exclusion reason: Does not report naturopathic consultations; |
| **Complementary and alternative health care in Israel** | Shuval, J. T.; Averbuch, E. | 2012 | Israel Journal of Health Policy Research | 1 | 1 | 7 | Exclusion reason: Does not report naturopathic consultations; |
| **WHO global report on traditional and complementary medicine 2019** | World Health Organisation | 2019 | | | | | Exclusion reason: Does not report naturopathic consultations; |
| **TRADITIONAL AND COMPLEMENTARY MEDICINE IN PRIMARY HEALTH CARE** | World Health Organisation | 2018 | | | WHO/HIS/SDS/2018.37 | | Exclusion reason: Does not report naturopathic consultations; |
| **The Philippines Health System Review** | World Health Organisation | 2018 | Health Systems in Transition | 8 | 2 | 352 | Exclusion reason: Does not report naturopathic consultations; |
| **SURGICAL WORKFORCE IN INDIA** | World Health Organisation | 2015 | | | | | Exclusion reason: Does not report naturopathic consultations; |
| **The prevalence and experience of Australian naturopaths and Western herbalists working within community pharmacies. B** | | 2011 | Journal of the Australian Traditional-Medicine Society | 17 | 3 | 167-168 | Exclusion reason: Duplicate; |
| **Use of traditional medicine and complementary and alternative medicine in Taiwan: a multilevel analysis** | Yeh, Mei-Ling; Lin, Kuan-Chia; Chen, Hsing-Hsia; Wang, Yu-Jen; Huang, Yu-Chiao | 2015 | Holistic Nursing Practice | 29 | 2 | 87-95 | Exclusion reason: Does not report naturopathic consultations; |
| **Benchmarks for training in traditional /complementary and alternative medicine: benchmarks for training in naturopathy** | World Health Organisation | 2010 | | | | | Exclusion reason: Not original research; |
| **Malaysia health system review** | World Health Organisation | 2012 | Health Systems in Transition | 2 | ISBN 978 92 9061 584 2 | 122 | Exclusion reason: Not original research; |

| | | | | | | | |
|---|---|---|---|---|---|---|---|
| **New Zealand health system review** | World Health Organisation | 2014 | Health Systems in Transition | 4 | | 272 | Exclusion reason: Not original research; |
| **The Regional Strategy for Traditional Medicine in the Western Pacific (2011â€“2020)** | World Health Organisation | 2012 | | | ISBN 978 92 9061 559 0 | 71 | Exclusion reason: Not original research; |
| **WHO traditional medicine strategy: 2014-2023.** | World Health Organisation | 2013 | | | | 78 | Exclusion reason: Not original research; |
| **Two-Thirds of Survey Respondents in Southern Sweden Used Complementary or Alternative Medicine in 2015** | Wemrell, M.; Merlo, J.; Mulinari, S.; Hornborg, A. C. | 2017 | Complementary medicine research | 24 | 5 | 302-309 | Exclusion reason: Does not report naturopathic consultations; |
| **Determinants for the Use of Complementary and Alternative Medicine: Results from a National Study** | Watts, Kristen Allen; Turner, Lori W. | 2018 | | | 10934635 | 307 | Exclusion reason: Does not report naturopathic consultations; |
| **Distribution of complementary and alternative medicine (CAM) providers in rural New South Wales, Australia: a step towards explaining high CAM use in rural health?** | Wardle, Jon; Adams, Jon; Magalhaes, Ricardo J. Soares; Sibbritt, David | 2011 | The Australian journal of rural health | 19 | 4 | 197-204 | Exclusion reason: Does not report naturopathic consultations; |
| **The interface with naturopathy in rural primary health care: A survey of referral practices of general practitioners in rural and regional New South Wales, Australia** | Wardle, J. L.; Sibbritt, D. W.; Adams, J. | 2014 | BMC Complementary and Alternative Medicine | 14 | | 238 | Exclusion reason: Does not report naturopathic consultations; |
| **Mapping the natural health landscape: New Zealand-based CAM professionals survey** | Vempati, R.; Dunn, J.; Cottingham, P.; Sibbritt, D.; Adams, J. | 2012 | BMC Complementary and Alternative Medicine | 12 | SUPPL. 1 | | Exclusion reason: Conference abstract only; |
| **Use of Complementary and Alternative Medicine in Bayamon, Puerto Rico** | Torres-Zeno, R. E.; Rios-Motta, R.; Rodriguez-Sanchez, Y.; Miranda-Massari, J. R.; Marin-Centeno, H. | 2016 | Puerto Rico Health Sciences Journal | 35 | 2 | 69-75 | Exclusion reason: Does not report naturopathic consultations; |
| **Attitude of Conventional and CAM Physicians Toward CAM in India** | Telles, Shirley; Gaur, Vaishali; Sharma, Sachin; Balkrishna, Acharya | 2011 | Journal of Alternative & Complementary Medicine | 17 | 11 | 1069-1073 | Exclusion reason: Does not report naturopathic consultations; |
| **Wellness versus treatment? Complementary and integrative healthcare (CIH) in the 2007 national health interview survey (NHIS)** | Stussman, B.; Alekel, L.; Nahin, R.; Edwards, E.; Barnes, P. | 2012 | BMC Complementary and Alternative Medicine | 12 | SUPPL. 1 | | Exclusion reason: Conference abstract only; |
| **Generational differences in complementary medicine use in young Australian women: Repeated cross-sectional dataset analysis from the Australian longitudinal study on women's health** | Steel, A.; Munk, N.; Wardle, J.; Adams, J.; Sibbritt, D.; Lauche, R. | 2019 | Complementary Therapies in Medicine | 43 | | 66-72 | Exclusion reason: Does not report naturopathic consultations; |
| **Complementary and alternative medicine: attitudes, knowledge and use among surgeons and anaesthesiologists in Hungary** | Soos, Sandor Arpad; Jeszenoi, Norbert; Darvas, Katalin; Harsanyi, Laszlo | 2016 | BMC Complementary and Alternative Medicine | 16 | 1 | 443 | Exclusion reason: Does not report |

| | | | | | | | |
|---|---|---|---|---|---|---|---|
| | | | | | | | naturopathic consultations; |
| **Complementary and alternative medicine: contemporary trends and issues** | Smith, Joanna M.; John Sullivan, S.; David Baxter, G. | 2011 | Physical Therapy Reviews | 16 | 2 | 91-95 | Exclusion reason: Not original research; |
| **Use of complementary and alternative medicine in the population of Kedah Darul Aman, Malaysia** | Sivadasan, S.; Ali, A. N.; Lin, L. W.; Balakrishnan, D.; Ramachandran, S.; Dhanaraj, S. A. | 2014 | International Journal of Pharmaceutical Sciences and Research | 5 | 4 | 1263-1273 | Exclusion reason: Does not report naturopathic consultations; |
| **Epidemiology of the use of complementary and alternative medicine in central area of Sao Paulo** | Simoes, O.; Castro, B. | 2013 | European Journal of Epidemiology | 28 | 1 SUPPL. 1 | S219 | Exclusion reason: Conference abstract only; |
| **[Complementary and alternative medicine services in Colombia]** | Rojas-Rojas, Alejandra | 2012 | Servicios de medicina alternativa en Colombia. | 14 | 3 | 470-7 | Exclusion reason: Does not report naturopathic consultations; |
| **Composition and distribution of the health workforce in India: estimates based on data from the National Sample Survey** | Rao, K. D.; Shahrawat, R.; Bhatnagar, A. | 2016 | WHO South-East Asia journal of public health | 5 | 2 | 133-140 | Exclusion reason: Does not report naturopathic consultations; |
| **Prevalence of Complementary and Alternative Medicine Use in the General Population in the Czech Republic** | Pokladnikova, J.; Selke-Krulichova, I. | 2016 | Forschende Komplementarmedizin (2006) | 23 | 1 | 22-28 | Exclusion reason: Does not report naturopathic consultations; |
| **Regional variation in use of complementary health approaches by U.S. adults** | Peregoy, J. A.; Clarke, T. C.; Jones, L. I.; Stussman, B. J.; Nahin, R. L. | 2014 | NCHS Data Brief | | 146 | 1-Aug | Exclusion reason: Does not report naturopathic consultations; |
| **Utilization of traditional and complementary medicine in Indonesia: Results of a national survey in 2014-15** | Pengpid, S.; Peltzer, K. | 2018 | Complementary Therapies in Clinical Practice | 33 | | 156-163 | Exclusion reason: Does not report naturopathic consultations; |
| **Complementary and alternative medicine (CAM) utilization in Texas hospices** | Olotu, B.; Brown, C. M.; Lawson, K.; Barner, J. C. | 2012 | Value in Health | 15 | 4 | A25 | Exclusion reason: Conference abstract only; |
| **Complementary and alternative medicine utilization in Texas hospices: Prevalence and challenges** | Olotu, B.; Brown, C.; Barner, J.; Lawson, K. | 2012 | Journal of the American Pharmacists Association | 52 | 2 | 215-216 | Exclusion reason: Conference abstract only; |
| **Experiences and meanings of integration of TCAM (Traditional, Complementary and Alternative Medical) providers in three Indian states: results from a cross-sectional, qualitative implementation research study** | Nambiar, D.; Narayan, V. V.; Josyula, L. K.; Porter, J. D. H.; Sathyanarayana, T. N.; Sheikh, K. | 2014 | BMJ Open | 4 | 11 | e005203 | Exclusion reason: Does not report naturopathic consultations; |
| **Naturopaths in Ontario, Canada: Geographic patterns in intermediately-sized metropolitan areas and integration implications** | Meyer, S. P. | 2017 | Journal of Complementary and Integrative Medicine | 14 | 1 | 92 | Exclusion reason: Does not report |

| | | | | | | | |
|---|---|---|---|---|---|---|---|
| | | | | | | | naturopathic consultations; |
| **An investigation into the use of complementary and alternative medicine in an urban general practice** | McKenna, F.; Killoury, F. | 2010 | Irish Medical Journal | 103 | 7 | | Exclusion reason: Does not report naturopathic consultations; |
| **A survey to explore the views and practices of CAM practitioners in the UK** | Majumdar, A.; Williams, S.; Adams, N. | 2012 | BMC Complementary and Alternative Medicine | 12 | SUPPL. 1 | | Exclusion reason: Conference abstract only; |
| **The prevalence of traditional and complementary medicine in the general population in Kashan, Iran, 2014** | Lotfi, M. S.; Adib-Hajbaghery, M.; Shahsavarloo, Z. R.; Gandomani, H. S. | 2016 | European Journal of Integrative Medicine | 8 | 5 | 661-669 | Exclusion reason: Does not report naturopathic consultations; |
| **Examining costs, utilization, and driving factors of complementary and alternative medicine (CAM) services** | Lewing, B.; Sansgiry, S. S. | 2018 | Value in Health | 21 | Supplement 1 | S97 | Exclusion reason: Conference abstract only; |
| **Profiling the Australian Consumer of Complementary and Alternative Medicine: A Secondary Analysis of National Health Survey Data** | Leach, M. J. | 2016 | Alternative therapies in health and medicine | 22 | 4 | 64-72 | Exclusion reason: Does not report naturopathic consultations; |
| **Complementary and alternative medicine (CAM) as part of primary health care in Germany-comparison of patients consulting general practitioners and CAM practitioners: A cross-sectional study** | Krug, K.; Kraus, K. I.; Herrmann, K.; Joos, S. | 2016 | BMC Complementary and Alternative Medicine | 16 | 1 | 409 | Exclusion reason: Does not report naturopathic consultations; |
| **Understanding CAM use in Lebanon: Findings from a national survey** | Kharroubi, S.; Chehab, R. F.; El-Baba, C.; Alameddine, M.; Naja, F. | 2018 | Evidence-based Complementary and Alternative Medicine | 2018 | | 4169159 | Exclusion reason: Does not report naturopathic consultations; |
| **Use of complementary and alternative medicine in Europe: Health-related and sociodemographic determinants** | Kemppainen, Laura M.; Kemppainen, Teemu T.; Reippainen, Jutta A.; Salmenniemi, Suvi T.; Vuolanto, Pia H. | 2018 | Scandinavian Journal of Public Health | 46 | 4 | 448-455 | Exclusion reason: Does not report naturopathic consultations; |
| **Complementary and alternative medicine usage in patients for different ailments in rural region of malwa area of punjab: A cross-sectional study** | Kaur, K.; Singh, B.; Kaur, G. | 2016 | National Journal of Physiology, Pharmacy and Pharmacology | 6 | 5 | 394-398 | Exclusion reason: Does not report naturopathic consultations; |
| **Determinants of patients preferring Complementary and Alternative medicine attending public hospitals in Lahore, Pakistan** | Hussain, A.; Ayesha,; Mufti, R. K.; Shahid, M.; Hassan, M. N.; Sultan, T.; Zahid, M. N.; Ali, I.; Iqbal, H. | 2018 | Journal of the Pakistan Medical Association | 68 | 6 | 914-918 | Exclusion reason: Does not report naturopathic consultations; |
| **State and Regional Comparisons of the Use of Complementary Health Approaches: National Health Interview Survey, 2012** | Jones, Lindsey; Peregoy, Jennifer; Stussman, Barbara; Nahin, Richard | 2014 | Journal of Alternative & Complementary Medicine | 20 | 5 | A143-A143 | Exclusion reason: Conference abstract only; |

| | | | | | | | |
|---|---|---|---|---|---|---|---|
| **Knowledge, attitude and practice of complementary and alternative medicine: A patient's perspective** | Jaiswal, K. M.; Bajait, C. S.; Pimpalkhute, S. A.; Dakhle, G. N.; Sontakke, S. D.; Magdum, A. | 2013 | Indian Journal of Pharmacology | 45 | SUPPL. 1 | S221 | Exclusion reason: Does not report naturopathic consultations; |
| **Use of complementary and alternative medicine within Norwegian hospitals** | Jacobsen, R.; FÃ¸nnebÃ¸, V. M.; Foss, N.; Kristoffersen, A. E. | 2015 | BMC Complementary & Alternative Medicine | 15 | 1 | 1-Jun | Exclusion reason: Does not report naturopathic consultations; |
| **Association between belief and attitude toward preference of complementary alternative medicine use** | Islahudin, F.; Shahdan, I. A.; Mohamad-Samuri, S. | 2017 | Patient Preference and Adherence | 11 | | 913-918 | Exclusion reason: Does not report naturopathic consultations; |
| **Patients' use of CAM: Results from the Health Survey for England 2005** | Hunt, K. J.; Ernst, E. | 2010 | Focus on Alternative and Complementary Therapies | 15 | 2 | 101-103 | Exclusion reason: Does not report naturopathic consultations; |
| **The utilization of complementary and alternative medicine in Taiwan: An internet survey using an adapted version of the international questionnaire (I-CAM-Q)** | Huang, C. W.; Tran, D. N. H.; Li, T. F.; Sasaki, Y.; Lee, J. A.; Lee, M. S.; Arai, I.; Motoo, Y.; Yukawa, K.; Tsutani, K.; Ko, S. G.; Hwang, S. J.; Chen, F. P. | 2019 | Journal of the Chinese Medical Association | 82 | 8 | 665-671 | Exclusion reason: Does not report naturopathic consultations; |
| **Utilization of complimentary and alternative health services in Iceland** | Helgadottir, B.; Vilhjalmsson, R.; Gunnarsdottir, T. J. | 2010 | Laeknabladid | 96 | 4 | 267-273 | Exclusion reason: Does not report naturopathic consultations; |
| **The use of complementary and alternative medicine in Iceland: Results from a national health survey** | Gunnarsdottir, T. J.; Orlygsdottir, B.; Vilhjalmsson, R. | 2019 | Scandinavian Journal of Public Health | | | 1.40 E+15 | Exclusion reason: Does not report naturopathic consultations; |
| **The Natural Medicine Workforce in Australia: A National Survey Part 1** | Grace, S.; Rogers, S.; Eddey, S. | 2013 | Journal of the Australian Traditional-Medicine Society | 19 | 1 | 13-18 | Exclusion reason: Does not report naturopathic consultations; |
| **The natural medicine workforce in Australia: A national survey Part 2** | Grace, S.; Rogers, S.; Eddey, S. | 2013 | Journal of the Australian Traditional-Medicine Society | 19 | 2 | 79-86 | Exclusion reason: Does not report naturopathic consultations; |
| **Complementary alternative medicine (CAM) use in Ireland: A secondary analysis of SLAN data** | Fox, P.; Coughlan, B.; Butler, M.; Kelleher, C. | 2010 | Complementary Therapies in Medicine | 18 | 2 | 95-103 | Exclusion reason: Does not report naturopathic consultations; |
| **Who uses complementary and alternative therapies in regional South Australia? Evidence from the Whyalla Intergenerational Study of Health** | D'Onise, K.; Haren, M. T.; Misan, G. M. H.; McDermott, R. A. | 2013 | Australian Health Review | 37 | 1 | 104-111 | Exclusion reason: Does not report naturopathic consultations; |

| | | | | | | | |
|---|---|---|---|---|---|---|---|
| **The characteristics, experiences and perceptions of naturopathic and herbal medicine practitioners: results from a national survey in New Zealand** | Cottingham, P.; Adams, J.; Vempati, R.; Dunn, J.; Sibbritt, D. | 2015 | Journal of the Australian Traditional-Medicine Society | 21 | 2 | 130-130 | Exclusion reason: Does not report naturopathic consultations; |
| **Integration of complementary and alternative medicine into medical schools in Austria, Germany and Switzerland - Results of a cross-sectional study** | Brinkhaus, B.; Witt, C. M.; Jena, S.; Bockelbrink, A.; Ortiz, M.; Willich, S. N. | 2011 | Wiener Medizinische Wochenschrift | 161 | 1-Feb | 32-43 | Exclusion reason: Does not report naturopathic consultations; |
| **The use of complementary therapies in Chile: Results from the national health survey 2010-2011** | Bedregal, P.; Passi, A.; Guerra, X.; Chang, M. | 2016 | Journal of Alternative and Complementary Medicine | 22 | 6 | A103-A104 | Exclusion reason: Conference abstract only; |
| **Complementary and Alternative Medicine (CAM) among adults in Italy: Use and related satisfaction** | Barbadoro, P.; Chiatti, C.; D'Errico, M. M.; Minelli, A.; Pennacchietti, L.; Ponzio, E.; Prospero, E. | 2011 | European Journal of Integrative Medicine | 3 | 4 | e319-e326 | Exclusion reason: Does not report naturopathic consultations; |
| **A preliminary study of complementary and alternative medicine (CAM) practitioners in Singapore** | Ang, S. C.; Wilkinson, J. M. | 2013 | Complementary Therapies in Medicine | 21 | 1 | 42-49 | Exclusion reason: Does not report naturopathic consultations; |
| **Use of complementary and alternative medicine among asthmatic patients in primary care clinics in Malaysia** | Alshagga, M. A.; Al-Dubai, S. A.; Muhamad Faiq, S. S.; Yusuf, A. A. | 2011 | Annals of Thoracic Medicine | 6 | 3 | 115-119 | Exclusion reason: Does not report naturopathic consultations; |
| **Knowledge, attitude and practice toward complementary and traditional medicine among Kashan health care staff, 2012** | Adib-Hajbaghery, M.; Hoseinian, M. | 2014 | Complementary Therapies in Medicine | 22 | 1 | 126-132 | Exclusion reason: Does not report naturopathic consultations; |
| **A survey of complementary and alternative medicine in Iran** | Abolhassani, Hassan; Naseri, Mohsen; Mahmoudzadeh, Sanam | 2012 | Chinese Journal of Integrative Medicine | 18 | 6 | 409-416 | Exclusion reason: Does not report naturopathic consultations; |
